# Short-term Air Pollution Exposure and Risk of Airway Inflammatory Response in Children (CHERISH): Protocol for a Randomised Mixed Factorial Study

**DOI:** 10.64898/2026.05.28.26353607

**Authors:** Scarlett Moloney, Hajar Hajmohammadi, Helen E Wood, Mohammed I Mead, Ian S Mudway, Gioia Mosler, Abigail C Thomson, Irene Gonzalez Calvo, James Scales, Abigail Whitehouse

## Abstract

**Introduction:** Air pollution is the largest environmental risk to human health. Children are disproportionately affected by air pollution and their exposure is amplified during physical activity. Observed concentrations of nitrogen dioxide in 1 in 4 London school playground exceeds the European limit, but the health impacts of air pollution exposure in London school playgrounds remain unexplored. Our study aims to assess and compare the acute changes in lung function and airway inflammation of primary school-aged children exercising in school playgrounds.

**Methods and analysis:** 330 children aged 8–11 years from ten London schools will be recruited to complete 90 minutes of physical activity and 90 minutes of rest in their school playground in a randomised crossover design. Pre-, post-, and 24-hour post-exposure oscillometry measurements will be performed with airway resistance at 5 Hz (R5) the primary physiological outcome. Nasal lavage samples will be collected pre-exposure and 24-hour post-exposure for analysis of inflammatory, oxidative, and vascular biomarkers, with IL-6 as the primary biological outcome. Mixed-effects regression models will examine associations between estimated pollutant exposures, exercise and physiological responses.

**Ethics and Dissemination:** This study has been approved by Queen Mary Ethics of Research Committee (QME25.1220). The study will be conducted according to the principles of the Helsinki Agreement (2013). Findings will be published in a peer-reviewed journal and shared at national and international conference presentations. This study will provide the first evidence on the short-term respiratory and immunological impacts of air pollution exposure during physical activity in London school playgrounds, supporting future public health guidance, school policy, and clinical advice for children, including those with asthma.

**Strengths and limitations of this study:**

- This study uses a randomised crossover design, allowing each child to act as their own control and reducing inter-individual variability in physiological responses.
- The study includes a large sample of 330 children from 10 London schools, providing sufficient statistical power and enabling assessment across a gradient of real-world air pollution exposures.
- Conducting the study during typical school playground activities enhances ecological validity and ensures findings are directly relevant to real-world school environments.
- As exposures occur in natural outdoor settings, air pollution levels and meteorological conditions cannot be fully controlled, which may introduce variability in exposure estimates.
- The study examines short-term physiological and immunological responses only, and therefore cannot determine the long-term health impacts of repeated exposure

## 1. Introduction

Air pollution is the fourth leading risk factor for global mortality [1]. Although the United Nations recognizes clean air as a human right, 99% of the global population breathes air that exceeds World Health Organization (WHO) guidelines for key pollutants, such as fine particulate matter (PM_2.5_) and nitrogen dioxide (NO_2_) (WHO, 2022). Long-term exposure to such pollutants is linked to increased risk of adverse health conditions across the lifespan including asthma [2], hypertension [3], cardiovascular disease [4] and dementia [5] with vulnerable groups, including the elderly children, at most risk. Children are disproportionately affected by air pollution due to their developing lungs and immune systems, higher ventilation rates relative to body mass, and greater time spent outdoors [6,7]. In London, air pollution levels in approximately one in four playgrounds exceed the UK annual legal limit of 40 μg/m^3^ for NO_2_ [8], with those in more deprived areas experiencing 8% higher concentrations than in the least deprived areas. Long-term exposure to NO_2_ above this limit has been associated with a 5% reduction in children’s lung function [9], and exposure to traffic-related air pollution (TRAP), including traffic-derived NO_2_ has been shown to impair lung development, exacerbate respiratory symptoms, and increase the risk of developing asthma [10,11]. Daily exposures to PM_2_._5_ and PM_10_ are also associated with acute decreases in children’s lung function indices with these associations strengthening after adjustment for NO_2_ [12]. Furthermore, paediatric asthma-related hospital admissions in England rose by 8% for every 10 μg/m^3^ increase in NO_2_ on the day of admission and during the four preceding days (five-day moving average) between 2011 and 2015 [13]. School playgrounds are central to children’s daily physical and social activities, serving as a hub for break times, physical education, and extracurricular sports. While physical activity provides substantial physical and mental health benefits [14–16], increased ventilation rate and depth during exercise, together with the switch from nasal to oral breathing, elevates pollutant uptake compared to rest. It has been reported that children living in areas with high ozone levels who participated in three or more team sports had a greater risk of developing asthma than their less active peers [17]. More recently, it is suggested that long-term higher outdoor activity levels were associated with a lower ratio of forced expiratory volume in 1 second to forced vital capacity (FEV_1_/FVC) [18]. However, no studies have compared the short-term effects of exposure to ambient air pollution during exercise versus rest on children’s lung and immune function.

Airway resistance at 5 and 20 Hz (R5-20) are indicators of small-airway obstruction and have been shown to be sensitive markers of air-pollution exposure in adults, with resistance increasing after exposure during physical activity [19]. Children experience over twice the deposited particle dose during physical activity due to their higher ventilation rates and smaller body mass [19], meaning the impacts on children are physiologically very different. Given greater exposure due to the amount of time spent in their school playground, and their physiological vulnerability, relying solely on data from adult participants may lead to an underestimation of the short-term effects of air pollution exposure during exercise on children’s health.

To date, the health impacts of air pollution exposure in school playgrounds remain unexplored. Therefore, the primary aim of this study is to assess and compare the acute health effects (changes in lung function and airway inflammation) of primary school-aged children exercising in school playgrounds with predicted contrasts in exposure to TRAP.

### 2. Methods and Analysis

### 2.1 Study Design

The CHERISH study will employ a randomised mixed factorial design with one within-subject factor (exposure condition: active vs. sedentary) and air pollution as a continuous variable. We will recruit 10 schools with different levels of air pollution, as measured by the Breathe Network [21]. An average of 33 participants will be recruited from each school according to a set of inclusion and exclusion criteria (presented in *Table 1*).

**Table 1.** CHERISH study inclusion and exclusion criteria.

| Inclusion Criteria | Exclusion Criteria |
| --- | --- |
| 1. Children aged 8 to 11<br>2. Attending Central and East London schools recruited to the study | 1. Not able to engage with PE lessons on safety grounds reported by their parents.<br>2. Children with learning or physical disabilities sufficient for them to be unable to give informed assent to the study, or to carry out study procedures. |

### 2.2 Aims/Outcomes

#### 2.2.1 Primary Aim

- To assess and compare the acute health effects (changes in lung function and airway inflammation) in children (aged 8-11 years) during active versus sedentary exposures in school playgrounds selected to maximise the contrast in traffic-derived pollutants, particularly NO_2_, within the study domain.

#### 2.2.2 Secondary Aims

- The provision of an air pollutant database (PM_2.5_ and PM_10_ mass, and NO_2_) covering all exposure days and one week prior to exposure days at participating school sites and children’s home addresses.
- To examine the relationship between variations in daily air quality and the physiologic and immunologic responses observed
- The provision of free outreach and engagement workshops for all schools, using Breathe London real-time, user-friendly air quality data to empower schools to monitor their own air quality.

#### 2.2.3 Outcome Measures

- Primary physiological: oscillometry (R5)
- Primary biological: Airway inflammation as measured by nasal mucosal immune responses (IL6) [22]
- Secondary: Nasal mucosal immune responses (Th1/Cellular Immunity: Granzyme A, Granzyme B; Acute/Innate Inflammation: IL-1β, IL-8, TNFα, MPO; Th2/Type 2 Immunity: IL-4, IL-5; Factors that promote the Th2 Response: TSLP and IL-33; Dual or Regulatory Role Mediators: IL-10; TH17-Associated: IL-23; Vascular/Endothelial Markers: VEGF, E-selectin; Oxidative stress: Glutathione, LOPs), Fractional Expired Nitric Oxide (FeNO), oscillometry (R5-20,).

### 2.3 Study Interventions

During two school visits, separated by a minimum two-week wash out period, children will take part in the following 90-minute school playground exposures in a randomised crossover design:

- Active: Multi-sport coaches from West Ham United Football Club Foundation will lead an educational outreach PE session designed to maintain a consistent level of exercise intensity, monitored by Actigraph physical activity monitors.
- Sedentary: Outreach scientists from QMULs Centre of the Cell will lead a ‘snotty science’ STEM science workshop explaining how air pollution impacts health.

The order in which the two exposures will take place will be randomised at the school level, by a blinded member of the research team via sealed envelopes containing one of the two conditions. The overall protocol, and the participant flow through the two separate exposures, are summarized in *Figure 1*.

**Figure 1.**
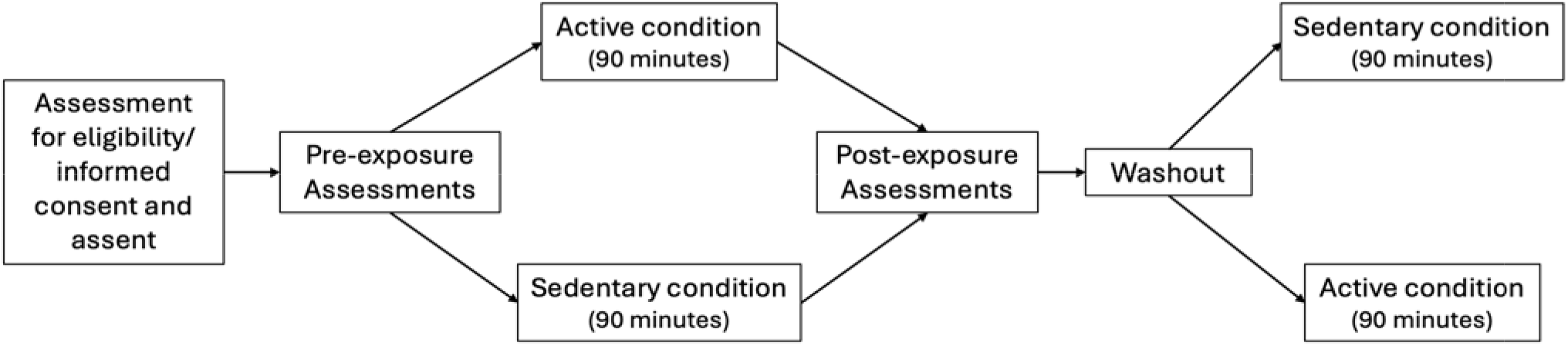
Diagram of participant involvement during the study.

Participants will perform identical nasal sampling and lung function assessments preceding and following each exposure (see assessment and sampling time points presented in *Table 2*). If children report having asthma or using an inhaler we shall also ask that they perform a FeNO test to quantify their immune status.

**Table 2.** Schedule of assessment and exposure.

|  | Pre-Study | Initial Assessment | Exposure | Post | 24-hour Post |
| --- | --- | --- | --- | --- | --- |
| Consent/Assent | X |  |  |  |  |
| Baseline Survey | X |  |  |  |  |
| Activity Monitor |  |  | X |  |  |
| FeNO |  | X |  |  |  |
| Oscillometry |  | X |  | X | X |
| Nasal Lavage |  | X |  |  | X |

#### 2.3.1 Sample Size

A total of ten schools will be purposively recruited using historical air quality data from Breathe London to include schools with varying levels of NO_2_ pollution. This strategy will provide the largest air pollution gradient possible between the school playgrounds. An average of 33 children will be recruited at each school, with an expected 10% drop.

For power calculation, we set significance level = 0.05, mean value = 0.572 and standard deviation = 0.14 for airway resistance (R5). If we predict a 5% change in R5, with total of 330 children (33 per school) and clustered within 10 schools (active vs sedentary), we will then have a power of 90.21%. Even if we have a dropout rate of 10 to 15%, we will have a power of 85.28%. 5% change in R5 was chosen as it is a clinically relevant change in airway restriction. This is especially true in the specific context of school playground exposures which occur daily in school environments.

#### 2.3.2 Exposure Site Selection

Central and East London schools will be selected from the Breathe London Network Communities to ensure a gradient in NO_2_ between the schools. The Breathe London Communities project have provided historical data which shows that we are able to provide the required air quality differences by working with schools already within their network. Planned study sites are shown in *Figure 2*.

**Figure 2.**
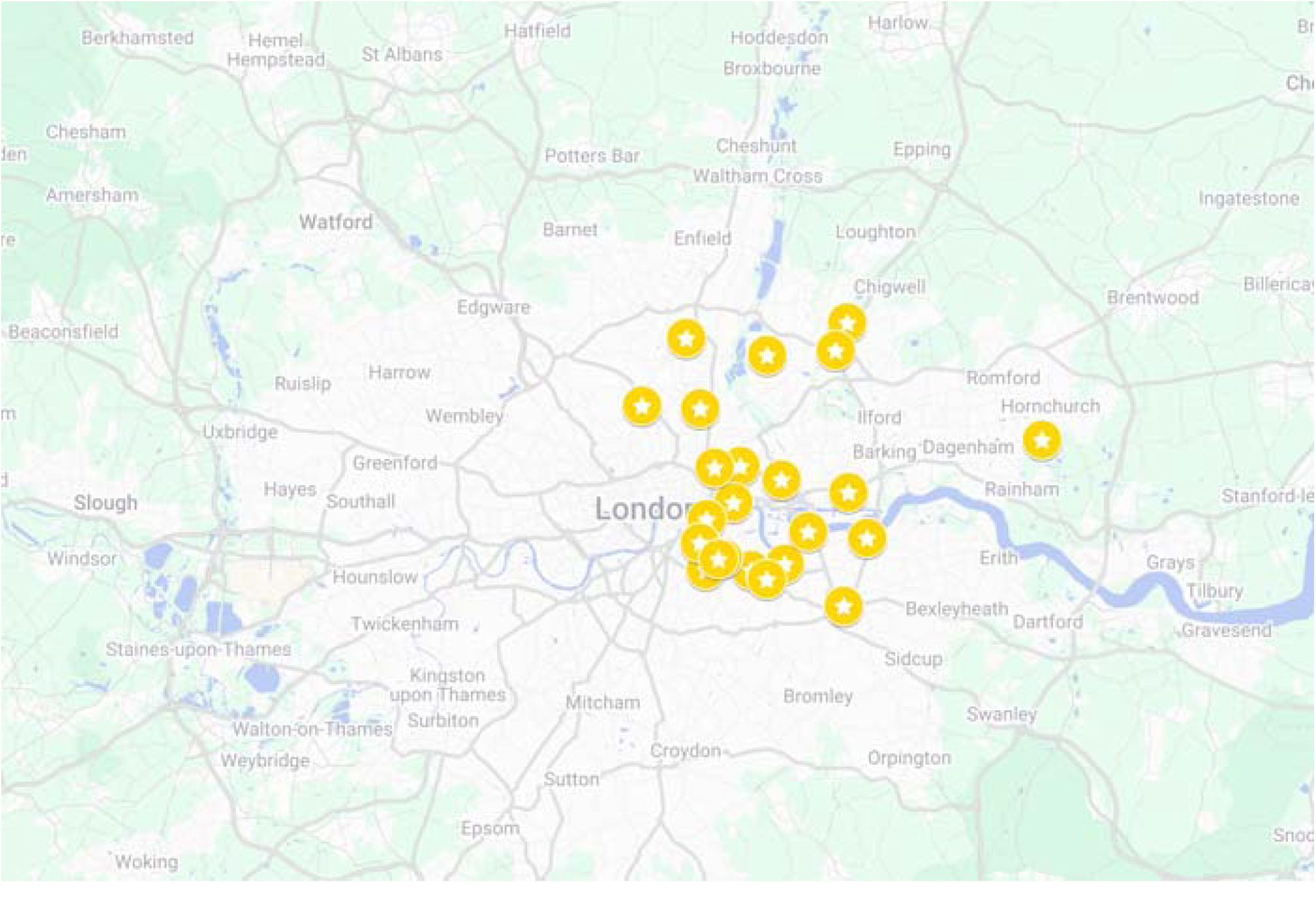
Map of targeted study sites.

#### 2.3.3 Recruitment

Participants will be recruited via a combination of letters sent home in school bags, and school and SMS systems. Information sheets, consent forms and parent questionnaires may be provided (and completed) electronically and/or on paper. Following their involvement, participants will be offered stickers and pens.

#### 2.3.4 Patient and Public Involvement

Both teachers and children were invited to give their input and feedback on the protocol design. As the study progresses, a patient and public involvement (PPI) group will be invited to project management group (PMG) meetings. The group will collaborate as advisors and partners in the study and are invited to provide unsolicited advice. To ensure the PPI group can provide meaningful advice and guidance, the CI will regularly update them on study developments via email. The meetings will follow the EUPATI (European Patients Academy on Therapeutic Innovation) roadmap (https://bit.ly/3bugCMk) discussing themes of: Research design and planning, research conduct, operations, dissemination and communication.

### 2.4 Exposure

To control for variations in air quality, exposures will take place between 10:30am and 13:30pm on weekdays. We will standardise exercise intensity to control physiological variation such as cardiovascular and ventilatory drift and to standardize inhaled doses of PM and gaseous pollutants over the 90-minute period and reflect a typical PE lesson. Moderate intensity was chosen, as prior research by the authors has shown that children spend break time performing moderate-to-vigorous levels of physical activity, ensuring a safe level of intensity, and reflecting children’s typical exercise levels and therefore ensuring maximum relevance of the findings. Regarding air pollution concentrations, there are currently no stipulated guidelines that restrict children from participating in outdoor activities or continuing their usual activities, irrespective of medical background. Therefore, data collection will proceed regardless of the air pollution concentrations. Exposure assessments will be conducted between November 2025 and May 2026.

While schools were recruited based on pollution exposure, they reflect the social and ethnic make-up of the wider east London area and so includes children from financially disadvantaged communities and ethnically diverse groups, including those who have English as an additional language, who often are not included in research studies. Air pollution exposure was estimated using data from the Breathe London air quality network, with NO_2_, PM_10_, and PM_2_._5_ concentrations derived from sensors located at participating school sites. Prior to school recruitment, historical data were extracted for the period October 2024 to February 2025 and restricted to typical school hours (09:00–15:00) to align with the planned data collection timeframe (Table 3). Nineteen school-based monitors within the study area were included. Observed air pollution concentrations were broadly consistent with expected levels for East London.

**Table 3.** Historical air quality at target school sites, October 2024–February 2025.

| Pollutant | Median (µg/m<sup>3</sup>) | Lower Quartile | Upper Quartile |
| --- | --- | --- | --- |
| NO<sub>2</sub> | 16.34 | 9.82 | 20.68 |
| PM<sub>2.5</sub> | 7.62 | 4.21 | 9.24 |

The mean and upper quartile concentrations for both pollutants were below the UK legal annual mean limits (40 μg/m^3^ for NO_2_ and 20 μg/m^3^ for PM_2_._5_). Median NO_2_ and PM_2_._5_ concentrations were also below the World Health Organization (WHO) guideline values for long-term exposure (10 μg/m^3^ for NO_2_ and 5 μg/m^3^ for PM_2_._5_). Consequently, pollution levels at participating schools are expected to comply with UK legal standards while at times exceeding WHO health-based recommendations.

### 2.5 Data collection before and after Exposure

Before and after each exposure, a battery of physiological measurements and samples will be taken to be assessed by the research team, which are appropriate for use in this population [23]. The time points of each measurement or sample are presented in an overview in *Figure 1*.

### 2.6 Respiratory Function Assessment

#### 2.6.1 Fractional Exhaled Nitric Oxide

The fractional exhaled nitric oxide (FeNO) can be an indicator of airway inflammation. FeNO will be measured using NIOX VERO (Niox, Oxford, UK) in accordance with the manufacturer’s instructions and the American Thoracic Society guidelines [24].

#### 2.6.2 Oscillometry

Airway resistance (R5, R20, and R5–20) will be measured using an oscillometer (Tremoflo® C-100, Thorasys, Montreal, QC, Canada) in accordance with manufacturer’s instructions and the European Respiratory Society guidelines [25].

### 2.7 Sample Collection

#### 2.7.1 Nasal Lavage

Nasal lavage samples will be collected using the nebuliser spray method [26] at two time points: pre-, and 24-hour post-exposure. The method consists of nebulizing 10 x 0.1 mL aliquots of Baxter’s 0.9% Sodium Chloride solution into each nostril and recovering the material into a sterile plastic receptacle. The procedure is repeated five times, with a total of 5 mL sprayed into each nostril. The samples are then stored at 4°C prior to transport to the laboratory facilities where they are centrifuged to remove mucus and cellular components at 800 × *g* at 4°C for 10 min (Thermo Scientific Heraeus Multifuge 3 Plus Centrifuge), prior to the cell-free supernatant being aliquoted for longer-term storage at ™80°C until required for analysis, or for longer-term storage as part of the sample biobank.

Samples will be assessed using a case-controlled study design. The cohort will consist of children who report asthmatic. We shall then randomly select a healthy comparator cohort, which is matched for 1) age 2) sex 3) Ethnicity 4) air pollution. Samples across the interval pre- to 24-hour post will be used to assess neutrophilic biology, oxidative stress pathways, and epithelial injury.

The primary hypothesis is that pollutant exposure will increase IL-6, a central cytokine involved in early inflammatory signaling and a key driver of Th17 polarisation. Secondary analyses will assess broader inflammatory pathways. Th1/cellular immunity will be evaluated through Granzyme A and Granzyme B, reflecting cytotoxic activation. Acute innate inflammatory responses will be captured via IL-1β, IL-8, TNF-α, and MPO, which are typically upregulated in response to particulate- and oxidant-induced airway injury. To characterise type-2 immune activation, we will measure IL-4 and IL-5, alongside epithelial-derived alarmins (TSLP, IL-33) that initiate and amplify Th2-skewing. IL-10 will be included as a mediator with dual anti-inflammatory or regulatory roles, while IL-23 will help define TH17-associated pathways linked to pollutant-induced immune shifts. To assess vascular involvement, VEGF and E-selectin will serve as indicators of endothelial activation. Finally, glutathione and lipid oxidation products (LOPs) will be quantified as markers of oxidative stress, a central mechanism by which short-term pollution exposure may trigger downstream immune effects. Overall, we hypothesise that higher acute pollutant exposure will induce a higher coordinated increase in inflammatory, oxidative, and vascular biomarkers consistent with acute immune activation.

## 3. Data Analysis

### 3.1 Statistical Methods

We will use a mixed effects regression model to determine the effects of the measured pollutants on changes in acute lung function and nasal lavage markers (as described in sections 2.6 and 2.7), defined as the difference in the paired measures between pre-exposure and post-exposure (at 0, 2, or 24-hours). For each endpoint, one key post-exposure time point will be selected based on the time of maximal effect observed in previous controlled exposure studies; for example, for R5 this will be 24-hour post-exposure.

We will quantify the effects of pollution exposure and physical activity (within participants). Concentrations (e.g. integrated average concentrations reported as interquartile ranges) of each pollutant will be included in the model. We will include pollutant as a continuous variable. The effects of different pollutants will be compared based on statistical significance and effect sizes calculated for selected exposure contrasts.

If sufficient variation exists, two-pollutant models will also be specified to evaluate which selected pollutant has the strongest association with each endpoint. In our models, we will account for potential confounders that may be present, as the exposure contrast cannot be entirely predicted by design. We may therefore include meteorological variables (e.g. temperature, relative humidity, wind speed and wind direction during the exposure) as well as individual characteristics such as age, sex, asthma status.

To account for repeated measures, random intercepts for participants will be included in the models, and random slopes will be explored if model convergence permits. Covariates will be included in the model to account for repeated measures (health outcome at baseline) and to adjust differences between groups (BMI as a Z-score, ethnicity, age, sex, asthma status). To account for possible clustering effects (children nested in schools) we shall first run null models to evaluate the interclass correlations (ICC) if the ICC effect is large at the school level we shall include two hierarchical levels by including school and student as random effects in the model. Any participants with missing data will be excluded.

All mixed models will be implemented in R (version 4.1.3 or newer; R Development Core Team), using functions from the *lme4* and *nlme* packages [27,28].

### 3.3 Governance

An Independent Steering Committee (ISC) chaired by Chris Griffiths will monitor and advise the study’s conduct and progress on behalf of the sponsor and funder. The ISC will meet with the Chief Investigator and study team three times during the two-year study. At least one member of the PPI group will be included in the meetings. The study is registered at clinicaltrials.gov (NCT07431021).

### 3.4 Consent

Parents will be required to provide written consent and participants to give written and verbal assent prior to any study procedures. Parents and participants will be given a minimum of 24 hours to consider their involvement and ask any questions. To ensure informed consent, further support will be provided in the form of playground information sessions, parent talks, school assemblies, school newsletters, and class talks - informing children and parents about the study and providing opportunity to ask questions.

### 3.5 Data Management

Data will be collected on paper or electronic consent forms and case report forms (CRFs)/questionnaires. All collected data will be held on backed-up encrypted servers. Any data transfers between ICL and QMUL will be performed via Secure File Transfer Protocols. Paper records, CRFs, and consent forms will be held locally in double-locked locations at QMUL.

### 3.6 Confidentiality

We will follow the best practice guidelines provided in Standard Operating Procedures (SOPs) by the Joint Research Management Office (JRMO) at QMUL. Paper records will be stored securely in locked filling cabinets in password-locked rooms in the pass-protected Centre for Primary Care. Electronic records will be stored in a password-protected study database on a secure server in the Centre for Primary Care. In the study, the database and personal details (name, address, date of birth) will be kept separate from the research data, which will be identified by a unique study reference number. In the tables of data, participants will only be identified by number, not by initials or name. Data management procedures will be completed in compliance with GDPR and trial regulations. Study data will be stored in the QMUL data safe haven, where data will be held in a UK server and access will be facilitated via two-factor authentication. All data will be backed up weekly to ensure that the data are safeguarded from accidental loss. The data will only be accessed and used by those members of the research team at QMUL and Imperial College London and the representatives of the sponsor who have been granted permission. Any required audits will be conducted by the Joint Research Management Office (JRMO).

### 3.7 Record Retention and Archiving

In accordance with the UK Policy Framework for Health and Social Care Research, research records will be kept for 20 years after the study has been completed, while personal records will be stored for one year after study completion. At the completion of the study, data will be moved to a trusted archive centre. At the end of the retention period, data will be destroyed in accordance with the best practice guidelines at the time of destruction.

### 3.8 Adverse Events Reporting

A risk and adverse events register will be maintained for the duration of the study. Adverse events will be discussed with the Chief Investigator and reported promptly to the sponsor as per Good Clinical Practice regulations and reviewed at study Project Management Group meetings.

### 3.9 Ethics and Dissemination

This study has been approved by Queen Mary Ethics of Research Committee (QME25.1220). The study will be conducted according to the principles of the Helsinki Agreement (2013).

Results arising from this study will be reported to the relevant funding agencies, scientific community, and partner groups through the following means:

1. Webinars on websites of our institutions to provide more detailed summaries of results, with downloads of key documents.
2. Presentations, especially to London organisations including the Greater London Authority (GLA), councils, school and local education authorities, and health and wellbeing boards.
3. Peer-reviewed publications.
4. Presentations at national and international conferences.

## 4. Discussion

Chronic exposures to air pollution have been shown to impair lung growth in primary school-aged children [29], however, the contribution of exercise to potentiating the adverse effects of air pollution on lung function are not well understood. Children are regularly exposed to elevated air pollution levels in school environments, with playgrounds identified as potential hotspots for short-term pollution peaks [30]. Exposure is further amplified during physical activity, when increased ventilation rates lead to greater inhaled doses of pollutants. The WHO recommends that children engage in at least 60 minutes of moderate-to-vigorous physical activity per day [31], much of which may occur in outdoor settings such as school playgrounds. A review of the limited existing literature on the association between air pollution, activity behaviour and health outcomes suggests that children who are active in polluted air may be at more risk of adverse health outcomes than their less active counterparts [32]. However, it remains unclear whether undertaking physical activity in polluted environments increases short-term health risks compared with sedentary behaviour.

Current public health and urban design strategies encourage active play and the provision of equipment and playground designs that promote movement and exercise [33]. Yet, despite increasing concerns about children’s exposure to air pollution in school settings, no studies have directly examined the short-term health effects of such exposure in London’s school playgrounds.

The UK Government’s Committee on the Medical Effects of Air Pollutants (COMEAP) identifies children with asthma as a susceptible group for adverse health outcomes from short-term pollution exposure. There is a critical evidence gap regarding how physical activity in polluted air affects these children, particularly whether the immediate physiological benefits of exercise outweigh the transient risks associated with increased pollutant uptake.

The CHERISH study will address this gap by being the first to investigate the acute health impacts of air pollution exposure among children in London’s school playgrounds. Findings from this research will inform local and city-wide policy, support the development of evidence-based guidelines for urban and school design, and help healthcare professionals provide more tailored advice to parents of children with respiratory conditions.

## Data Availability

This manuscript describes a study protocol and no individual participant data have yet been generated. The historical air pollution data referred to in the manuscript were obtained from the Breathe London air quality monitoring network and are publicly accessible through Breathe London. De-identified participant-level data generated during the CHERISH study may be made available on reasonable request after study completion, subject to ethical approval, participant consent, data governance requirements, and appropriate data-sharing agreements.

https://www.breathelondon.org/

## Author Contributions

JS, AW, HEW and HH conceived the project. All authors were involved in the practical aspects of project delivery. JS, AW, HEW and HH developed the study analysis plan. All authors contributed to the drafting of the paper. All authors have read and agreed to the published version of the manuscript.

## Acknowledgements

The authors would like to thank our Independent Scientific Committee, and particularly Prof Klea Katsouyanni for her input and guidance

## Disclaimer/Publisher’s Note

The statements, opinions and data contained in all publications are solely those of the individual author(s) and contributor(s) and not of MDPI and/or the editor(s). MDPI and/or the editor(s) disclaim responsibility for any injury to people or property resulting from any ideas, methods, instructions or products referred to in the content.

## List of Abbreviations

ATS: American Thoracic Society
BMI: Body Mass Index
CI: Chief Investigator
COMEAP: Committee on the Medical Effects of Air Pollutants
CRF: Case Report Form
ERS: European Respiratory Society
EUPATI: European Patients’ Academy on Therapeutic Innovation
FEV_1_: Forced Expiratory Volume in 1 Second
FVC: Forced Vital Capacity
FeNO: Fractional Exhaled Nitric Oxide
GDPR: General Data Protection Regulation
GLA: Greater London Authority
IL: Interleukin (e.g., IL-1β, IL-4, IL-5, IL-6, IL-8, IL-10, IL-23, IL-33)
ISC: Independent Steering Committee
JRMO: Joint Research Management Office
LOPs: Lipid Oxidation Products
MPO: Myeloperoxidase
NO_2_: Nitrogen Dioxide
PE: Physical Education
PIS: Participant Information Sheet
PNC: Particle Number Concentration
PM_2.5_: Particulate Matter ≤2.5 μm
PM_10_: Particulate Matter ≤10 μm
PMG: Project Management Group
PPI: Patient and Public Involvement
R5–R20: Measures of Airway Resistance at 5 and 20 Hz
SOP: Standard Operating Procedure
STEM: Science, Technology, Engineering & Mathematics
TNF-α: Tumour Necrosis Factor Alpha
TRAP: Traffic-Related Air Pollution
TSLP: Thymic Stromal Lymphopoietin
UK: United Kingdom
UN: United Nations
VEGF: Vascular Endothelial Growth Factor
WHO: World Health Organization

## Appendices

1. **Supplementary Materials 1:** Model consent form and other related documentation given to participants – add the CRFs and PISs
2. **Supplementary Materials 2:** Physical Activity lesson plans

