## Supplementary 1 for "Short-term Air Pollution Exposure and Risk of Airway Inflammatory Response in Children (CHERISH): Protocol for a Randomised Mixed Factorial Study"

CHERISH
Children's Health, Respiratory Inflammation and Short-term air pollution

### Information Sheet for Parents

### The Children's Health Respiratory Inflammation and Short-term Air Pollution (CHERISH) Study

**Queen Mary Ethics of Research Committee reference number:** QME25.1220

#### Summary

We are doing a study to find out how London's air pollution affects children's health during PE lessons. We want to find out:

- How pollution affects lung function (breathing) and immune response (how the body fights infection).
- How the body defends itself from pollution, using nasal mucus (snot) samples.

This sheet explains why we are doing the study and what it involves. Please read it carefully. Talk to others if you wish. You do not have to give a reason if you decide you don't want you child to take part.

##### What is the purpose of the study?

- Air pollution from traffic is harmful, especially for children as their lungs are still growing.
- One in four London school playgrounds have air pollution levels above the legal limit.
- We want to find out if playing outside in this pollution harms children's health.

##### What will taking part involve?

- Our team will visit the school twice.
- Coaches from West Ham Football Club's charity will run a 90-minute PE lesson
- Our outreach scientists will also run a 90-minute snotty science lesson.
- Before the lesson:
  - Your child will do a breathing test to check their lung health:
  - They will wear an activity monitor during the lesson.
  - They will provide a snot sample by squirting salty water up their nose and letting it drip into a cup.
- After the lesson:
  - They will repeat the breathing test and snot sample immediately and again the next day.
- All this will take place during a normal double period lesson.
- We will also ask if your child has asthma, breathing symptoms or uses an inhaler.
- If your child has asthma, we would ask them to do another breathing test (FeNO) which will help us understand the type of asthma they have.

Breathing test video

##### Why is my child being invited?

- They are being invited because their school has agreed to take part in the study.
- Your child should not take part if you do not let them take part typical PE lessons (such as limited mobility or severe illness).



Barts and The London School of Medicine and Dentistry
IMPERIAL
WEST HAM UNITED LONDON FOUNDATION
CARRii Transforming Lung Health. Together. Centre of Applied Respiratory Research Innovation and Impact

#### Does my child have to take part?

No, it is up to you to decide whether they should take part. If you decide they can take part, you will need to complete either the paper or online consent form. Your child can choose not to take part in this study or stop taking part in it if they change their mind, at any time and without giving a reason. If you do decide they can take part you will be free to withdraw your child from the study at any time without needing to provide a reason, and with no penalties or detrimental effects. The team will ask your child if they are happy to take part before both sessions.

#### What are the possible benefits of my child taking part?

- Your child will get a free health check of their lungs and if we find anything wrong, we can provide you with a letter to take to their GP, explaining what we have found.
- The findings from this research will be used to inform government air quality policy.

#### What are the possible disadvantages and risks of taking part?

- There is very little risk.
- All tests are commonly used and are safe.
- The PE lessons will follow the national curriculum.
- There is a risk that physical activity may instigate a flare up if your child has underlying respiratory conditions.
- There is a chance that we may identify medical conditions through the health assessment. We would notify you if this happens.

#### What information will be collected about my child?

- Ethnicity, as this affects lung function results.
- Your home address, to estimate air pollution exposure before the visit.
- Their lung function scores, height and weight will also be recorded.
- Height and weight are used to estimate lung function
- We will collect samples of their snot
- We will also ask short survey about their respiratory health and exposure to air pollution

#### How will my child's data be stored and who will have access to it?

Any information we collect will be transferred to, stored and analysed at Queen Mary University of London. The data will be stored in de-identified format, with your child's name replaced by a unique code. To reduce the risk of disclosure, personal identifiers will be stored separately from the research data on secure computers. Any paper records of data (e.g. forms completed by the study team at school visits) will be stored either in locked filing cabinets within lockable offices, in an access-restricted building, or will be sent off site to a secure storage facility that complies with our security requirements. All data handling, processing, transfer and storage procedures comply with our obligations under General Data Protection Regulations and comply with our local data handling and security policies and procedures. Personal data will only be accessed and used by those members of the research team at QMUL and representatives of the sponsor who have been granted permission.

#### When and how will my child's data be destroyed?

Their data will be destroyed five years after the completion of the study (November 2032). All data will be destroyed in accordance with best practice at the time of destruction.

##### How will my child's data be used and shared?

We will publish the results in scientific journals and present them at research conferences (no names or individual data will be shown). We may place anonymised data in a shareable online data folder to support future research. Other researchers will need to request access through Dr James Scales and Dr Abi Whitehouse.

##### How will my child's snot samples be used?

- Samples will be stored securely at Imperial College London.
- They will be used to study how pollution affects lung health.
- Only research led by Dr James Scales and Dr Abi Whitehouse will use these samples.

##### Under what legal basis are you collecting this information?

- Queen Mary University of London processes personal data for research purposes in accordance with the lawful basis of 'public task'.
- Please read Queen Mary's privacy notice for research participants containing important information about your personal data and your rights in this respect.
- If you have any questions relating to data protection, please contact Queen Mary's Data Protection Officer, Queens' Building, Mile End Road, London, E1 4NS or or 020 7882 7596.

##### What will happen if I want to withdraw my child from this study?

- You can withdraw their participation in the study at any time without providing a reason.
- You can choose to withdraw from future participation but allow the data already collected to be used by the research team.
- Alternatively, you may also choose to withdraw any data already collected.
- You can choose to have the data destroyed at any time up to two months after the last day of their participation.

##### What should I do if I have concerns about this study?

- If you have any concerns about the manner in which the study was conducted, in the first instance, please contact the researchers responsible for the study: Dr James Scales and Dr Abi Whitehouse.
- If you have a complaint, which you feel you cannot discuss with the researchers then you should contact the Research Ethics Facilitators by.
- When contacting the Research Ethics Facilitators, please provide details of the study title (CHERISH), description of the study and QMERC reference number QMERC25.1220, the researchers involved, and details of the complaint you wish to make.

##### Who can I contact if I have any questions about this study?

- Dr James Scales
- Dr Abi Whitehouse
- CHERISH study team

### Children's Information Sheet

### (CHERISH) Children's Health, Respiratory Inflammation and Short-term Air Pollution

**Queen Mary Ethics of Research Committee reference number:** QME25.1220

**What is this study about?**

- We want to see how air pollution affects children's breathing during PE lessons.
- We also want to learn how your nose protects you from pollution.
- We will be working with many schools and children like you across London.

**What will happen if I take part?**

- Our team will come to your school twice.
- One time, you will have a 90-minute PE lesson.
- Another time you will have a snotty science lesson.

**Before the lessons:**

- You will do a breathing test by blowing into a tube.
- You will wear a small activity monitor.
- You will provide a snot sample by squirting salty water up your nose and letting it drip into a cup.

**After the lessons and the next day:**

- You will do the breathing test and snot sample again

##### What is good about taking part?

- You will get a check of your lungs to see how healthy they are.
- You will help scientists learn how to keep children healthy.

##### Do I have to take part?

- No. It is your choice.
- You can say no if you do not want to take part.
- You can change your mind any time.

##### Will it hurt?

- No, it will not hurt.
- The tests are safe, and hundreds of children have done them before.

##### What if I have questions or feel worried?

- You can talk to your teacher.
- You can talk to your parents or carers.
- You can talk to the study team when they visit.

**Thank you for reading!** We look forward to seeing you!

We start with a breathing and snot test.

Then will measure your height and weight.

You then have a PE lesson and a science lesson in a random order.

Finally, you do the tests again and we compare them.

Study ID Number: | | | – | 0 | | |

CHERISH
Children's Health, Respiratory Inflammation and Short-term air pollution

### Consent Form

#### (CHERISH) Children's Health, Respiratory Inflammation and Short-term Air Pollution

**Queen Mary Ethics of Research Committee reference number:** QME25.1220

**PLEASE SIGN EACH BOX TO SHOW YOU HAVE READ AND AGREE WITH THE FOLLOWING AND SIGN AND DATE AT THE BOTTOM OF THE PAGE:**

***PLEASE SIGN THESE BOXES***

1. I confirm I have read the parent information sheet (V2.0 23.10.2025) for the above study. I have had the opportunity to consider the information, ask questions and have had these answered satisfactorily. ☐
2. I understand that my child's participation is voluntary and that my child is free to withdraw at any time, without giving a reason, and without their care or legal rights being affected. ☐
3. I agree for my child to take part in health checks at their school, comprising measurement of height, weight, lung function, and immune response. ☐
4. I agree for my child to take part in the PE lesson and science workshop. ☐
5. I understand that Queen Mary University of London will use information about my child in order to undertake this study and will act as the data controller for this study. ☐
6. I understand that the information collected about my child may be shared completely anonymously with other researchers to support future non-commercial research *(optional)*. ☐
7. I understand I may be contacted in future to consider further health assessment studies of my child *(optional)*. ☐
8. I agree for my child to take part in the above study. ☐

If you have any questions relating to data protection, please contact Data Protection Officer, Queens' Building, Mile End Road, London, E1 4NS or or 020 7882 7596.

*WRITE **YOUR** NAME, DATE AND SIGN HERE:*

*Please **sign** in this box:*

Name of Parent or legal guardian ______ Date ______ Parent/guardian's Signature ______

*WRITE **YOUR CHILD'S** NAME, DATE AND **ASK THEM** TO SIGN HERE:*

*Ask **your child to sign** in this box:*

Name of Participant (child) ______ Date ______ Child's Signature ______

*LEAVE THIS FOR THE **RESEARCHER** TO COMPLETE:*

Name of person taking consent ______ Date ______ Signature ______

Study ID Number: [ ][ ] – 0 [ ][ ]
Date of completion: [ ][ ] – [ ][ ] – [ ][ ][ ][ ]
D D - M M - Y Y Y Y

### Case Report Form for Parents

### The Children's Health Respiratory Inflammation and Short-term Air Pollution (CHERISH) Study

It's great that you've decided your child can take part in the CHERISH research study. We need to ask you for some information about your child to help us to do the research.

Please complete the questions in this form as fully and accurately as you can

#### SECTION A: ABOUT YOUR CHILD

**First name:** ……………………………………..

**Last name:** ………………………………………………………………………..

**Date of Birth:** [ ][ ] – [ ][ ] – [ ][ ][ ][ ] (e.g. 01-JAN-2025)
D D - M M - Y Y Y Y

**Sex at birth:** (please tick **one** box)

- Male ☐
- Female ☐

**Ethnicity:** (please tick **one** box)

**Asian or Asian British**

- Indian ☐
- Pakistani ☐
- Bangladeshi ☐
- Any other Asian background ☐

**Black or Black British**

- Caribbean ☐
- African ☐
- Any other Black background ☐

**Mixed**

- White and Black Caribbean ☐
- White and Black African ☐
- White and Asian ☐
- Any other mixed background ☐

**White**

- British ☐
- Irish ☐
- Any other White background ☐

Study ID Number: [ ][ ]–[0][ ][ ]

**Chinese or any other ethnic group**

Chinese [ ]

Any other ethnic group [ ]

If Other, please specify: ………………………………………….

**Current home address** (this is used to estimate exposure to air pollution over the past 12 months):

House number/name and street: ..…………………………………………………………

Town/City: ……………………………………………………

Postcode: ……………………… (e.g. E1 2AB)

Has your child lived at this address for **less than 12 months**? (please tick **one**) Yes [ ] No [ ]

If **YES**, what date have they lived here from? [ ][ ]–[ ][ ]–[ ][ ][ ][ ] (e.g. 01-JAN-2025)

D D - M M - Y Y Y Y

And what was their **previous** home address?

House number/name and street: ………………...…………………………………………

Town/City: ……………………………………………………

Postcode: ……………………… (e.g. E1 2AB)

Country (if not UK): ………………………………………....

Start date: [ ][ ]–[ ][ ][ ]–[ ][ ][ ][ ] (e.g. 01-JAN-2018)

End date: [ ][ ]–[ ][ ][ ]–[ ][ ][ ][ ] (e.g. 01-JAN-2018)

D D - M M M - Y Y Y Y

#### SECTION B: ABOUT YOU

**Your name:** ……………………………………………….……………

**Your relationship to the child:** ……………………………………………………(e.g. mother, father)

We would like to keep in touch with you about the study, for example, to let you know about the study results and any further follow up. Your contact details will not be shared with anyone outside the study research team.

**Email address:** ……………………………………………………………………………………………

**Mobile phone number:** ……………………………………… (11 digits starting 07)

Study ID Number: ☐☐ – 0☐☐

#### SECTION C: Questionnaire

##### 1. Smoking

Does anyone who lives at home with your child smoke cigarettes? Yes ☐ No ☐

If **YES**, what is their relationship to the child? (e.g. mother, father, uncle)

…………………………………………………………

Does anyone who lives at home with your child use a vape/e-cigarette? Yes ☐ No ☐

If **YES**, what is their relationship to the child? (e.g. mother, father, uncle)

…………………………………………………………

##### 2. Indoor environment

What is the **main type of heating** you use in your home? (please tick **one**)

Gas boiler ☐
Gas heater ☐
Electric boiler ☐
Electric heater ☐
Heat pump ☐
District or community ☐
Other (please specify): ☐

**Do you use the following cooking appliances?** (please answer for each appliance)

Gas oven ☐
Gas hob ☐
Electric oven ☐
Electric hob ☐
Induction hob ☐
Microwave ☐
Air fryer ☐

Study ID Number: [ ][ ]-[0][ ][ ]

**Are you currently experiencing any of the following in your home?** (tick all boxes that apply)

Condensation ☐
Lack of adequate heating ☐
Draught ☐
Damp walls and/or floors ☐
Visible mould ☐
Dry air ☐
Damp air ☐
Stuffy air ☐
Unpleasant odour ☐
Excessive dust ☐
Pests (e.g cockroaches, mice, rats, moths, beetles, bed bugs, etc) - please specify: ☐

_ _ _ _ _ _ _ _ _ _ _ _ _ _ _ _ _ _ _ _ _ _ _ _ _ _ _ _ _ _ _ _ _ _ _ _ _ _ _ _ _ _ _ _ _ _ _ _ _ _ _ _ _ _ _ _

**Do you have any pets in your household**?

No ☐
Dog ☐
Cat ☐
Other – please specify: ☐

_ _ _ _ _ _ _ _ _ _ _ _ _ _ _ _ _ _ _ _ _ _ _ _ _ _ _ _ _ _ _ _ _ _ _ _ _ _ _ _ _ _ _ _ _ _ _ _ _ _ _ _ _ _ _ _

#### 3. Your child's breathing and other symptoms (ISAAC Questionnaire)

PLEASE COMPLETE THIS SECTION ABOUT **YOUR CHILD**

**Section 1: Core questionnaire for asthma**

1 Has your child <u>ever</u> had wheezing or whistling in the chest at any time in the past?

Yes ☐
No ☐

IF YOU HAVE ANSWERED "NO" PLEASE SKIP TO QUESTION 6

2 Has your child had wheezing or whistling in the chest <u>in the past 12 months?</u>

Yes ☐
No ☐

IF YOU HAVE ANSWERED "NO" PLEASE SKIP TO QUESTION 6

Study ID Number: [ ][ ] – [0][ ][ ]

---

3 How many attacks of wheezing has your child had <u>in the past 12 months?</u>

- None ☐
- 1 to 3 ☐
- 4 to 12 ☐
- More than 12 ☐

4 <u>In the past 12 months</u>, how often, on average, has your child's sleep been disturbed due to wheezing?

- Never woken with wheezing ☐
- Less than one night per week ☐
- One or more nights per week ☐

5 <u>In the past 12 months</u>, has wheezing ever been severe enough to limit your child's speech to only one or two words at a time between breaths?

- Yes ☐
- No ☐

---

6 Has your child <u>ever</u> had asthma?

- Yes ☐
- No ☐

7 <u>In the past 12 months</u>, has your child's chest sounded wheezy during or after exercise?

- Yes ☐
- No ☐

8 <u>In the past 12 months</u>, has your child had a dry cough at night, apart from a cough associated with a cold or chest infection?

- Yes ☐
- No ☐

**Section 2: Core questionnaire for rhinitis**

1 Has your child <u>ever</u> had a problem with sneezing, or a runny, or blocked nose when he/she DID NOT have a cold or the flu?

- Yes ☐
- No ☐

IF YOU HAVE ANSWERED "NO" PLEASE SKIP TO QUESTION 6

---

2 <u>In the past 12 months,</u> has your child had a problem with sneezing, or a runny, or blocked nose when he/she DID NOT have a cold or the flu?

- Yes ☐
- No ☐

IF YOU HAVE ANSWERED "NO" PLEASE SKIP TO QUESTION 6

Study ID Number: ☐☐ – 0 ☐☐

---

3 In the past 12 months, has this nose problem been accompanied by itchy-watery eyes?

Yes ☐
No ☐

4 In which of the past 12 months did this nose problem occur? (Please tick any which apply)

| | | | | | |
|---|---|---|---|---|---|
| January | ☐ | May | ☐ | September | ☐ |
| February | ☐ | June | ☐ | October | ☐ |
| March | ☐ | July | ☐ | November | ☐ |
| April | ☐ | August | ☐ | December | ☐ |

5 In the past 12 months, how much did this nose problem interfere with your child's daily activities?:

Not at all ☐
A little ☐
A moderate amount ☐
A lot ☐

---

6 Has your child ever had hayfever?

Yes ☐
No ☐

**Section 3: Core questionnaire for eczema**

1 Has your child ever had an itchy rash which was coming and going for at least six months?

Yes ☐
No ☐

IF YOU HAVE ANSWERED "NO" PLEASE SKIP TO QUESTION 7

---

2 Has your child had this itchy rash at any time in the past 12 months?

Yes ☐
No ☐

IF YOU HAVE ANSWERED "NO" PLEASE SKIP TO QUESTION 6

---

3 Has this itchy rash at any time affected any of the following places:

Yes ☐
No ☐

Study ID Number: | | | – | 0 | | |

the folds of the elbows, behind the knees,
in front of the ankles, under the buttocks,
or around the neck, ears or eyes?

4 At what age did this itchy rash first occur?

- Under 2 years ☐
- Age 2-4 years ☐
- Age 5 or more ☐

5 Has this rash cleared completely at any time during the past 12 months?

- Yes ☐
- No ☐

6 In the past 12 months, how often, on average, has your child been kept awake at night by this itchy rash?

- Never in the past 12 months ☐
- Less than one night per week ☐
- One or more nights per week ☐

---

7 Has your child ever had eczema?

- Yes ☐
- No ☐

**Thank you for taking the time to complete this form**

**Please contact us if you have any questions about this form or the CHERISH Study**


**CHERISH STUDY TEAM**

**Website:** (tbc)

**Principal Investigators:** Dr Abi Whitehouse & Dr James Scales

Study ID Number: | | | – | 0 | | |

### Case Report Form for Researchers:

### (CHERISH) Children's Health, Respiratory Inflammation and Short-term Air Pollution

#### PARTICIPANT INFORMATION

First name: …………………………………… Last name: ……………………………………

Date of birth: [ ][ ] – [ ][ ] – [ ][ ][ ][ ]
D D - M M - Y Y Y Y

School name: ………………………………………………………………………………………..

Year group: (tick **one**)

Year 4 ☐
Year 5 ☐
Year 6 ☐

**Forms received:**

**Signed and dated consent form** ☐
**Parent CRF** ☐

#### SCHOOL VISITS

| | **FIRST** | **SECOND** |
|---|---|---|
| Visit date (DD-MM-YYYY) | [ ][ ] – [ ][ ] – [ ][ ][ ][ ] | [ ][ ] – [ ][ ] – [ ][ ][ ][ ] |
| Participant outcome (exposure day) | Participated ☐<br>Absent ☐<br>Declined ☐<br>Withdrawn ☐ | Participated ☐<br>Absent ☐<br>Declined ☐<br>Withdrawn ☐ |
| Exposure | Physical activity ☐<br>Sedentary science lesson ☐ | Physical activity ☐<br>Sedentary science lesson ☐ |
| Participant outcome (24-hr post) | Participated ☐<br>Absent ☐<br>Declined ☐<br>Withdrawn ☐ | Participated ☐<br>Absent ☐<br>Declined ☐<br>Withdrawn ☐ |

Study ID Number: | | | – | 0 | | |

#### FIRST SCHOOL VISIT

| Assessment | Completed? |
|---|---|
| Participant assent and questionnaire | |
| Oscillometry (pre-exposure) | |
| Nasal lavage | |
| FeNO | |
| Height and weight measurement | |
| Physical activity monitor | |
| Exposure session | |
| Oscillometry (post-exposure) | |

**NOTES (e.g., regarding any completes tasks and/or reason for non-completion)**

……………………………………………………………………………………………………
……………………………………………………………………………………………………
……………………………………………………………………………………………………
……………………………………………………………………………………………………

#### ASSENT (read questions to the child)

Do you understand what we are doing today, or do you have any questions?

**ANSWER QUESTIONS/EXPLAIN AS REQUIRED**

Are you feeling well today? (please tick **one**)

Yes ☐
No ☐
Not sure ☐

If **No** or **Not Sure**, please explain:

……………………………………………………………………………………………………
……………………………………………………………………………………………………

Are you happy to take part in the study today? (please tick **one**)

Yes ☐
No ☐

**IF NO, DO NOT PROCEED (clarify whether child is declining on this occasion or wants to withdraw from the study)**

Study ID Number: | | | – | 0 | | |

#### QUESTIONNAIRE (read questions to the child)

How did you travel to school today? (please tick **all that apply**)

| | | | |
|---|---|---|---|
| Walk | ☐ | Taxi | ☐ |
| Scooter | ☐ | Bus | ☐ |
| Bike | ☐ | Train/Tube | ☐ |
| Private car | ☐ | Other | ☐ |

How do you usually travel to school?" (please tick **all that apply**)

| | | | |
|---|---|---|---|
| Walk | ☐ | Taxi | ☐ |
| Scooter | ☐ | Bus | ☐ |
| Bike | ☐ | Train/Tube | ☐ |
| Private car | ☐ | Other | ☐ |

Do you have asthma? (please tick **one**)

Yes ☐
No ☐
Don't know ☐

Do you have an asthma pump? (please tick **one**)
**(SHOW PICTURE, IF REQUIRED)**

Yes ☐
No ☐
Don't know ☐

**IF YES**

What colour pump do you have?
(please tick **all that apply**)

Blue ☐
Brown ☐
Purple ☐
Other ☐
Don't know ☐

Have you used your pump today?
(please tick **one**)

Yes ☐
No ☐
Don't know ☐

**IF YES**

Which colour pump have you used today?
(please tick **all that apply**)

Blue ☐
Brown ☐
Purple ☐
Other ☐
Don't know ☐

**IF YES, please ask for further details (time taken, how many pumps):**

**…………………………………………………………………………………………………………….**

Study ID Number: ☐☐-0☐☐

#### HEALTH ASSESSMENT

Height: ☐☐☐.☐ (cm)

Weight: ☐☐☐.☐ (kg)

FeNO: ☐☐☐ (ppb)

#### FIRST 24-hr POST VISIT

| **Assessment** | **Completed?** |
|---|---|
| Participant assent | |
| Oscillometry | |
| Lavage | |

#### ASSENT (read questions to the child)

Do you understand what we are doing today, or do you have any questions?

**ANSWER QUESTIONS/EXPLAIN AS REQUIRED**

Are you feeling well today? Yes ☐
(please tick **one**) No ☐
Not sure ☐

If **No** or **Not Sure**, please explain:

……………………………………………………………………………………………………………………

……………………………………………………………………………………………………………………

Are you happy to take part in the study today? Yes ☐
(please tick **one**) No ☐

**IF NO, DO NOT PROCEED (clarify whether child is declining on this occasion or wants to withdraw from the study)**

Study ID Number: | | |–|0| | |

#### SECOND SCHOOL VISIT

| Assessment | Completed? |
|---|---|
| Participant assent and questionnaire | |
| Oscillometry (pre-exposure) | |
| Nasal lavage | |
| FeNO | |
| Height and weight measurement | |
| Physical activity monitor | |
| Exposure session | |
| Oscillometry (post-exposure) | |

**NOTES (e.g., regarding any completes tasks and/or reason for non-completion)**

……………………………………………………………………………………………………

……………………………………………………………………………………………………

……………………………………………………………………………………………………

……………………………………………………………………………………………………

#### ASSENT (read questions to the child)

Do you understand what we are doing today, or do you have any questions?

**ANSWER QUESTIONS/EXPLAIN AS REQUIRED**

Are you feeling well today? (please tick **one**)

Yes ☐
No ☐
Not sure ☐

If **No** or **Not Sure**, please explain:

……………………………………………………………………………………………………

……………………………………………………………………………………………………

Are you happy to take part in the study today? (please tick **one**)

Yes ☐
No ☐

**IF NO, DO NOT PROCEED (clarify whether child is declining on this occasion or wants to withdraw from the study)**

Study ID Number: | | | – | 0 | | |

#### QUESTIONNAIRE (read questions to the child)

How did you travel to school today? (please tick **all that apply**)

| | | | |
|---|---|---|---|
| Walk | ☐ | Taxi | ☐ |
| Scooter | ☐ | Bus | ☐ |
| Bike | ☐ | Train/Tube | ☐ |
| Private car | ☐ | Other | ☐ |

How do you usually travel to school?" (please tick **all that apply**)

| | | | |
|---|---|---|---|
| Walk | ☐ | Taxi | ☐ |
| Scooter | ☐ | Bus | ☐ |
| Bike | ☐ | Train/Tube | ☐ |
| Private car | ☐ | Other | ☐ |

Do you have asthma? (please tick **one**)

Yes ☐
No ☐
Don't know ☐

Do you have an asthma pump? (please tick **one**)
**(SHOW PICTURE, IF REQUIRED)**

Yes ☐
No ☐
Don't know ☐

**IF YES**

What colour pump do you have?
(please tick **all that apply**)

Blue ☐
Brown ☐
Purple ☐
Other ☐
Don't know ☐

Have you used your pump today?
(please tick **one**)

Yes ☐
No ☐
Don't know ☐

**IF YES**

Which colour pump have you used today?
(please tick **all that apply**)

Blue ☐
Brown ☐
Purple ☐
Other ☐
Don't know ☐

**IF YES, please ask for further details (time taken, how many pumps):**

……………………………………………………………………………………………………….

Study ID Number: ☐☐-0☐☐

#### HEALTH ASSESSMENT

Height: ☐☐☐.☐ (cm)

Weight: ☐☐☐.☐ (kg)

FeNO: ☐☐☐ (ppb)

#### SECOND 24-hr POST VISIT

| **Assessment** | **Completed?** |
|---|---|
| Participant assent | |
| Oscillometry | |
| Lavage | |

#### ASSENT (read questions to the child)

Do you understand what we are doing today, or do you have any questions?

**ANSWER QUESTIONS/EXPLAIN AS REQUIRED**

Are you feeling well today? (please tick **one**)

- Yes ☐
- No ☐
- Not sure ☐

If **No** or **Not Sure**, please explain:

……………………………………………………………………………………………………………………

……………………………………………………………………………………………………………………

Are you happy to take part in the study today? (please tick **one**)

- Yes ☐
- No ☐

**IF NO, DO NOT PROCEED (clarify whether child is declining on this occasion or wants to withdraw from the study)**
